# Malaria Pre-screening Technology Using Artificial Intelligence (AI)

**DOI:** 10.64898/2026.07.15.26357432

**Authors:** Olive O. Ibeto, Ephraim O. Nwoye

## Abstract

Malaria remains a severe health problem in endemic regions because people lack adequate diagnostic tools, leading to delayed medical care and elevated death rates. This research introduces a dual-mode artificial intelligence system that uses two complementary models to enhance malaria pre-screening and diagnosis. The patient-centered model uses multivariate logistic regression to analyze biosignals, including heart rate, body temperature, and oxygen saturation, collected through a wearable sensor prototype and a mobile interface for symptom analysis. The system enables patients to begin self-assessment to determine their level of need before scheduling a doctor’s appointment. The clinician-centered model represents a customized convolutional neural network that uses annotated microscopy images of red blood cells to achieve 94.84% accuracy, 95.71% precision, 93.87% recall, 94.78% F1 score, and 0.84 Area Under Curve (AUC). The patient model achieved 94.6% accuracy and an AUC of 0.985 using a 70/30 train-test split. These systems work together to create a layered diagnostic system that can operate independently or together to detect malaria at an early stage, especially in areas with limited resources. The findings demonstrate that wearable biosignal data integration with image-based deep learning can produce dependable, scalable, and user-friendly systems for malaria pre-screening.

## 1. Introduction

Malaria exists as a global health emergency because it leads to fatal outcomes in areas without sufficient medical facilities and diagnostic equipment. In 2021, the global count of reported malaria cases reached 247 million in total, with a staggering 95% of these cases originating from the African continent. Among the registered cases, a significant 96% resulted in fatalities attributable to malaria. Notably, Nigeria accounted for a concerning 27% of these deaths, reflecting a high mortality rate within the country [1]. These statistics reveal that there is a critical public health challenge that persists, despite global interventions and already existing preventive measures. The gold standard for managing this disease uses laboratory diagnostic methods, which include Microscopy tests and Rapid Diagnostic Tests. These methods work well in clinical research environments, but they do not perform well in settings with limited resources because they face multiple barriers to fast diagnosis and treatment. The current limitations have the potential to generate adverse effects on malaria control programs, which affect resource-limited areas that require immediate intervention for their underserved communities.

The prevention of malaria requires immediate action because it stands as the main solution to decrease malaria-related deaths. The achievement of this goal depends on monitoring systems that enable early disease detection. Monitoring helps to keep track of the occurrence of a change in the normal levels of fluids or substances in the body. A change in the body levels can be a response to stimuli and can also indicate an impending illness. With advancements in technology, Artificial Intelligence (AI) has emerged as a powerful tool in controlling malaria. Its integration and advancement in healthcare have revolutionized the entire healthcare system and led to increased efficiency, reduced hospital stays, faster result analysis, and, most importantly, ultimately reducing the mortality rates across several diseases, including malaria.

AI has been successfully used for the prediction of disease [2], the development of models [3], and healthcare data analysis [4,5]. Moreover, non-invasive technology has emerged as a valuable approach in diagnosing, monitoring, and treating illnesses without the need for invasive procedures or devices that penetrate the body’s internal parts. This cost-effective technique has become instrumental in experimental settings and holds significant importance in the field of healthcare [6]. Irrespective of these advancements, most AI-driven malaria diagnostic approaches often have a narrowed focus, which relies on a single diagnostic pathway, such as image classification, vital signs, or symptom screening. This singular approach has the potential to limit effectiveness and generalizability

This study, therefore, seeks to develop an AI-based diagnostic framework for malaria detection, one that combines multiple diagnostic modalities. Specifically, we adopted a dual-mode framework that uses a patient-centered and a clinician-centered approach to malaria detection. The patient-centered approach uses biosignals (vital signs) associated with malaria, such as temperature, heart rate, oxygen saturation, and a mobile application for clinically-based symptom assessment, allowing users to input symptoms for AI-driven prescreening, while the clinician-centered arm focuses on medical image-based analysis using an AI model trained to detect malaria parasites in blood smear images, thereby improving diagnostic accuracy to confirm the malaria diagnosis.

The dual-pathway system improves diagnostic precision through its dual system, which enables quick identification and provides medical services to patients and healthcare providers. The system uses existing AI capabilities to create an efficient, non-invasive malaria detection system that provides accessible malaria diagnosis for low-resource settings to decrease undiagnosed infections and enhance health outcomes in these areas.

## 2. Literature Review

### 2.1 AI in Malaria Detection

The implementation of AI technology in malaria diagnosis systems helps solve problems that exist with current conventional diagnostic approaches. Specifically, machine learning algorithms have shown promising results in diagnosing malaria using clinical and microscopic data [7]. A comparison of various machine learning models for malaria diagnosis found that Random Forest performed best on a parasite-specific dataset regardless of class-balancing, while on a broader, more heterogeneous dataset, Random Forest achieved the best performance only after applying the Synthetic Minority Oversampling Technique (SMOTE) to address class imbalance [36]. Similarly, Yadav et al. demonstrated that Artificial Neural Networks and Support Vector Machines performed better than traditional rapid diagnostic tests, achieving over 92% [8]. Beyond just diagnosis alone, AI has also been applied to malaria epidemiology, with R programming used to build a dashboard supporting malaria surveillance and control [9]. However, the majority of research is concentrated on individual AI methods and sometimes lacks field implementation, particularly in low-resource environments.

### 2.2 Convolutional Neural Network (CNN) - Based Image Analysis For Malaria Detection

CNNs have successfully improved the diagnosis of malaria by enhancing image-based recognition of the parasite. For instance, Turuk et al. tested pre-trained CNN models such as AlexNet, ResNet50, and VGG19. They observed that VGG19 had the highest accuracy of 93.89% [10]. Additionally, Hasan-Sifat et al. demonstrated how AI can revolutionize the diagnosis of malaria. By leveraging U-Net for cell segmentation (97.67% accuracy), CNN for recognizing contaminated red blood cells (100% accuracy), and VGG16 for species classification (95.55% accuracy), showing high precision [11]. These findings demonstrate how AI can improve early detection, lower human error, and offer scalable solutions, especially in environments with limited resources. To further improve the CNN performance, Abdul Qayyum et al. introduced a robust CNN model enhanced with kernel dilation techniques, where the Fibonacci series-wise dilated CNN emerged as the top performer, achieving impressive metrics of 96.05% accuracy, 95.80% precision, 96.33% recall, and 96.06% F1-score on a large dataset of 27,558 cell images [12].

While traditional CNN models like VGG19 and ResNet50 are powerful, they require significant computational resources. Magotra & Rohil brought to the limelight the use of a lightweight CNN model that requires less computing power as compared to other heavier models like VGG-19 and Inception v3. Their model also achieved an impressive 96% accuracy [13]. This makes the model promising and perfect for resource-limited settings where every second counts. Similarly, the research of Goni et al. presented an explainable ultra-lightweight CNN that reached more than 99% accuracy in all evaluation metrics to demonstrate its effectiveness for malaria detection in restricted resource settings [34]. Building on this approach, Al Muazzaz and Goni developed a lightweight CNN model that reached 99.56% accuracy and processed images at 2 ms per prediction while demonstrating exceptional suitability for early malaria detection in resource-limited environments [35].

The new developments demonstrate how CNN-based methods operate as essential tools that boost malaria detection speed and accuracy while developing accessible solutions for areas with limited resources. It is also important to investigate other diagnostic methods, such as biosignal processing, in order to improve both the range and precision of malaria detection systems.

### 2.3 Biosignal Processing For Malaria Diagnosis

Malaria diagnosis often relies on biosignals, which are simple and non-invasive physiological signs. These signs are monitored using medical devices and systems. The monitoring system tracks vital signs through SpO_2_ readings, body temperature measurements, ECG data, and heart rate monitoring. They are essential because they show early changes in the body’s normal functioning when the infection begins to set in. Malaria infections start by causing fever and elevated heart rate before patients show any other noticeable symptoms. The early warning system delivers vital information that healthcare providers need to use during resource-limited situations when laboratory tests become unavailable. Medical staff can provide quick treatment that leads to better survival outcomes because they can track biosignals using basic, reliable monitoring systems. One study by Li et al. demonstrated how integrating signal processing techniques like transimpedance amplifiers (TIA) and Butterworth filters into Magnetoresistive (MR) sensor systems resulted in a low-noise, low-power, and highly sensitive diagnostic approach for malaria [14].

Signal processing isn’t only about monitoring vital signs. It plays a huge role in medical imaging, a critical piece in malaria diagnosis. Once an image has been obtained (usually a blood smear), it is analyzed effectively to spot the parasites. In medical image analysis, signal processing techniques have been applied for preprocessing, image enhancement, feature extraction, pattern recognition, and integration with deep learning. Ruiz-Alzola et al. highlighted the critical role of advanced signal processing techniques in biomedical imaging, emphasizing their application in improving image reconstruction, analysis, and interpretation across various modalities, including Magnetic Resonance Imaging (MRI), Electroencephalogram (EEG), and X-ray, to enhance clinical diagnostics and disease detection [15].

In the specific context of malaria blood smear analysis, preprocessing methods such as Contrast Limited Adaptive Histogram Equalization (CLAHE) and normalization have proven to be useful for improving the visibility of key features [16,17,18]. These techniques help make subtle details in stained smears stand out, so that AI or human examiners can spot parasites more easily. For feature extraction, methods like time-frequency analysis, the Fourier transform, and wavelet transform analyze spectral characteristics or features of images, capturing details like texture and morphology [19, 20,21]. The Sobel and Canny methods serve as edge detection algorithms that help identify image boundaries and structures for successful image segmentation and classification [22, 23, 24].

Together, these signal processing techniques can improve the accuracy and reliability of AI-powered diagnostic systems. These tools convert unprocessed images and body signals into useful medical data, which doctors can depend on during situations where they lack access to sophisticated laboratory facilities. The individual diagnostic methods, which include imaging, biosignals, and symptom assessment, provide important information, yet using one data source at a time creates gaps in diagnostic certainty. Researchers now use multimodal diagnostic systems, which unite multiple diagnostic methods into a single system to solve this problem.

### 2.4 Multi-Modal Approaches To Malaria Diagnosis

Multimodal methods unite clinical symptoms with medical imaging and biosignals to improve diagnostic accuracy and overcome the limitations of using single diagnostic approaches. The combination of different diagnostic methods produces a complete malaria infection evaluation, which enables prompt treatment and minimizes the risk of incorrect diagnosis.

The first step of symptom-based assessment functions as a screening method that identifies potential cases through patient-reported symptoms and doctor-observable clinical indicators [25]. Meanwhile, biosignals such as heart rate variability, oxygen saturation, and temperature readings provide non-invasive physiological markers that can indicate infection before symptoms develop and become visible [26]. In parallel, AI-powered image analysis of stained blood smears has demonstrated a high accuracy in detecting malaria parasites [33], further strengthening the diagnostic processes.

Despite their promise, multi-modal malaria diagnostic systems face several challenges. Data integration across diverse inputs, clinical validation in real-world settings, cost, and accessibility of wearable sensors can be barriers in a low-resource environment. Further research and development are needed to refine these approaches and confirm their effectiveness at scale.

## 3. Methods

This study’s methodology comprises two complementary systems: a patient-centered system and a clinician-centered system. The patient-centered system was designed using vital signs, clinical symptom assessment, and a multivariate logistic regression model to generate a probabilistic risk estimate, while the clinician-centered system employs a custom convolutional neural network (CNN) for image-based detection to support clinicians in malaria diagnosis. The datasets for both systems were obtained from publicly available malaria datasets on Kaggle.

### A. Patient-Centered System

This system was designed as an AI-driven predictive framework for malaria risk estimation based on symptom-related clinical variables. The model was developed using 309 records comprising sociodemographic attributes, vital signs, and symptom-related variables relevant to the study objectives. This dataset was originally obtained from a publicly available Kaggle repository and is provided as supplementary material accompanying this manuscript. Feature engineering and preprocessing were performed in R, including data cleaning, normalization, and selection of relevant predictor variables for model development. A multivariate logistic regression model was built (Table 1), with temperature, heart rate, and number of symptoms identified as the key predictive features. The dataset was split into 70% training and 30% testing sets, with ten-fold cross-validation applied to guard against overfitting during evaluation. Additional train-test splits (80/20, 90/10, 60/40, and 50/50) were also evaluated to assess model stability across partition ratios.

**Table 1.**
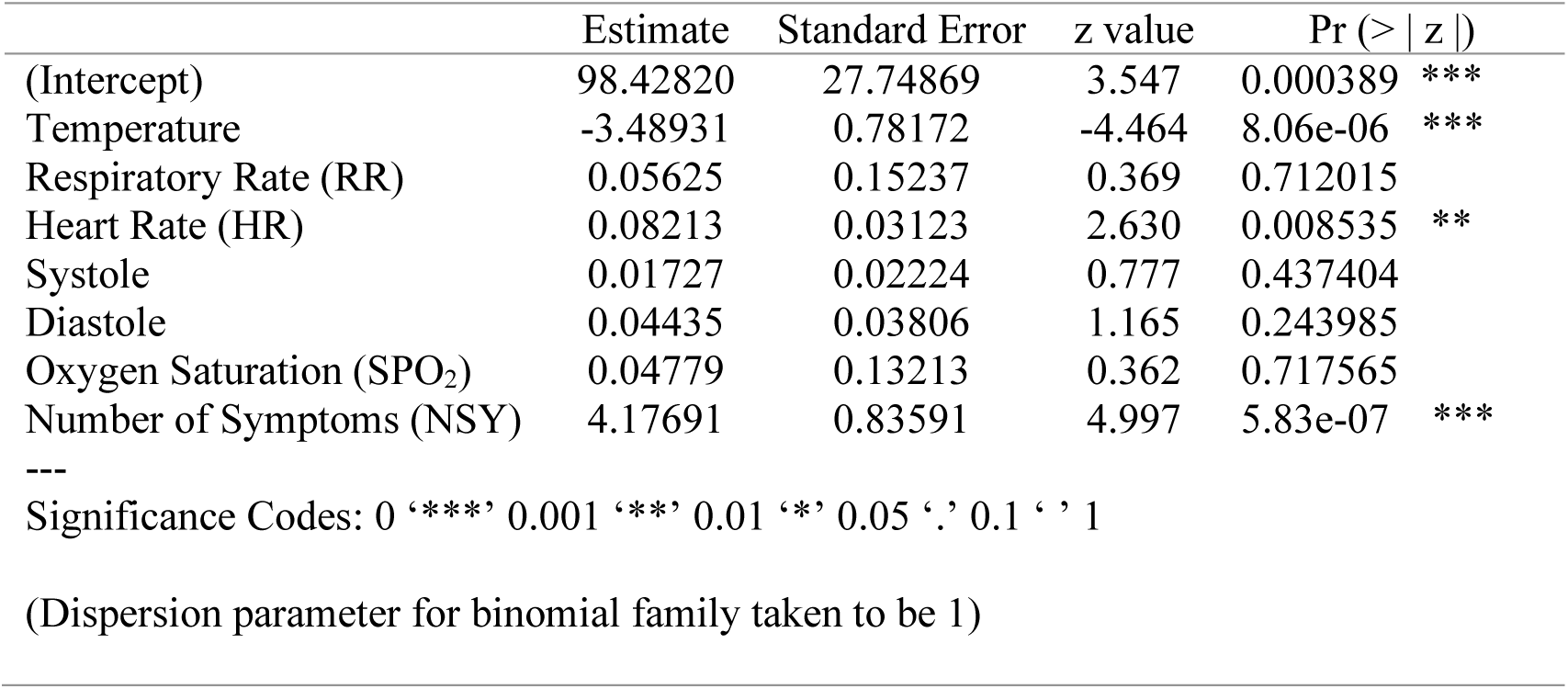
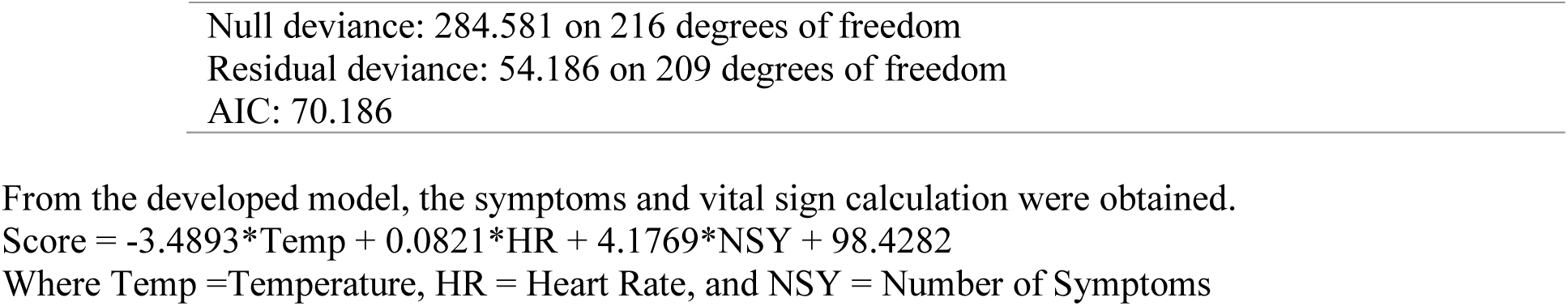
Coefficients of the Logistic Regression Model for Malaria Detection.

|  | Estimate | Standard Error | z value | Pr (> \|z\|) |
| --- | --- | --- | --- | --- |
| (Intercept) | 98.42820 | 27.74869 | 3.547 | 0.000389 *** |
| Temperature | -3.48931 | 0.78172 | -4.464 | 8.06e-06 *** |
| Respiratory Rate (RR) | 0.05625 | 0.15237 | 0.369 | 0.712015 |
| Heart Rate (HR) | 0.08213 | 0.03123 | 2.630 | 0.008535 ** |
| Systole | 0.01727 | 0.02224 | 0.777 | 0.437404 |
| Diastole | 0.04435 | 0.03806 | 1.165 | 0.243985 |
| Oxygen Saturation (SPO<sub>2</sub>) | 0.04779 | 0.13213 | 0.362 | 0.717565 |
| Number of Symptoms (NSY) | 4.17691 | 0.83591 | 4.997 | 5.83e-07 *** |

Significance Codes: 0 '\*\*\*' 0.001 '\*\*' 0.01 '\*' 0.05 '.' 0.1 ' ' 1
(Dispersion parameter for binomial family taken to be 1)
Null deviance: 284.581 on 216 degrees of freedom Residual deviance: 54.186 on 209 degrees of freedom AIC: 70.186

This score is used by the mobile application, Malacare, to analyze the patient’s symptoms and vital signs. Malacare was developed using MIT App Inventor and Java, with a connection to the Blynk database for data storage and exchange. The app features login authentication, a patient screening survey, and a regression model that classifies patients based on entered data. A binary classification system determines whether a patient is likely to have malaria, while an integrated wearable sensor provides real-time vital sign monitoring through WiFi.

This system provides patients with the unique opportunity to screen themselves for malaria symptoms, supporting early detection and reducing hospital burden, while also enabling clinicians to access structured patient data for informed decision-making. The system was designed to assist, rather than replace medical judgment. Fig. 1 illustrates the communication structure between the wearable sensor device and Malacare, which was developed using Proteus.

**Fig. 1.**
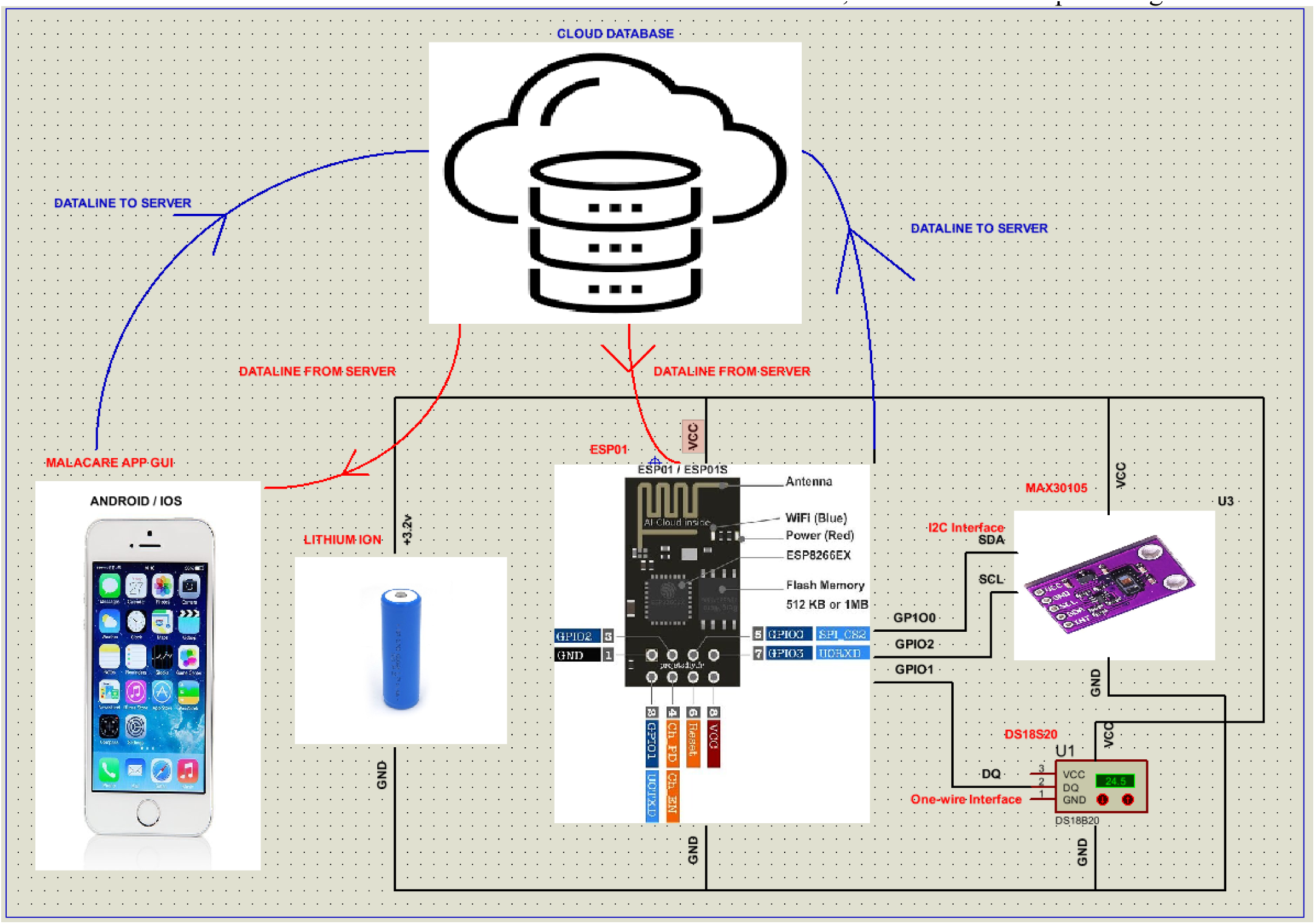
Wearable sensor device communication with Malacare

**Fig. 2.**
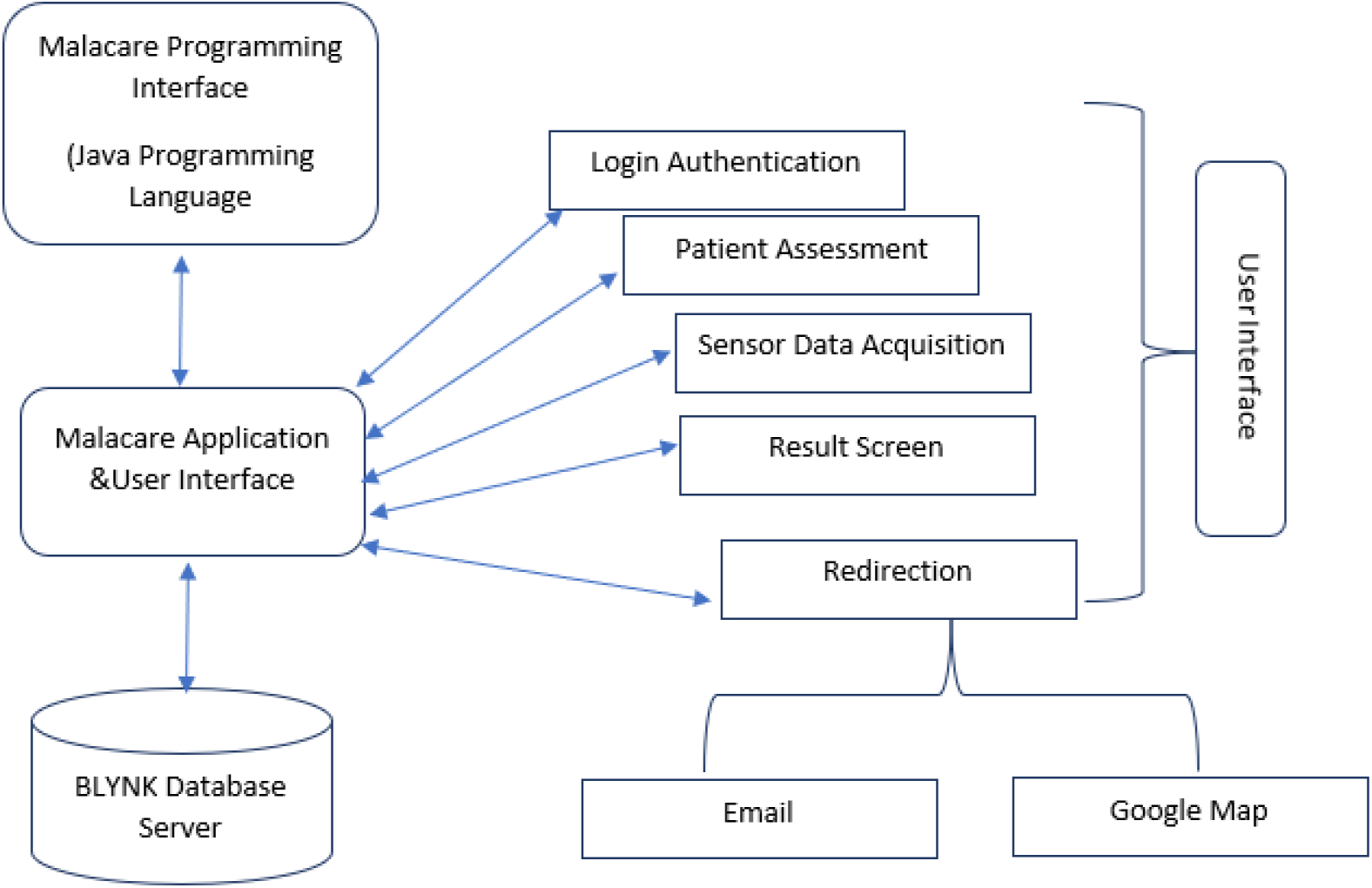
Mobile Application Structure

### B. Clinician-Centered System

The Clinician-centered system was developed to assist clinicians in malaria diagnosis by automating the analysis of microscopic images using deep learning. A custom CNN was designed and trained to classify parasitized and uninfected cells. The system was developed using Python, which incorporated machine learning libraries like TensorFlow and Keras machine learning libraries for model development, training, and optimization. The research dataset included 27,558 cell images, which were divided into 13,779 parasitized and 13,779 uninfected samples. The images underwent preprocessing, which included converting them to greyscale and resizing them to 224 x 224 pixels and normalizing pixel values for better and faster computational performance. A train-test split of 80/20 was used. A summary of the preprocessing is shown in Table 2.

**Table 2.** Data Preprocessing Summary.

| Step | Details | Value/Figure |
| --- | --- | --- |
| Image Resizing | Resized to 224x224 pixels | 224x224 pixels |
| Normalization | Pixel values scaled to range [0, 1] | Scaled to [0,1] |
| Data Split | 80-20 split between training and test sets | Training: 80%, Test: 20% |
| Labels Encoding | One-hot encoded labels for classification | 2 classes (Parasitized, Uninfected) |

The model architecture included multiple convolutional layers with increasing filters (32, 64, 128) for feature extraction. The next layer, which was the max-pooling layer, was used to reduce dimensionality, while the fully connected layers were used for classification. The final layer used the softmax activation function to categorize the images into two classes: Parasitized or uninfected.

Adam Optimizer was used to train the model with a learning rate of 0.001, using categorical cross-entropy as the loss function. Early stopping was also used to prevent overfitting. The CNN model Architecture is shown in Fig. 3

**Fig. 3.**
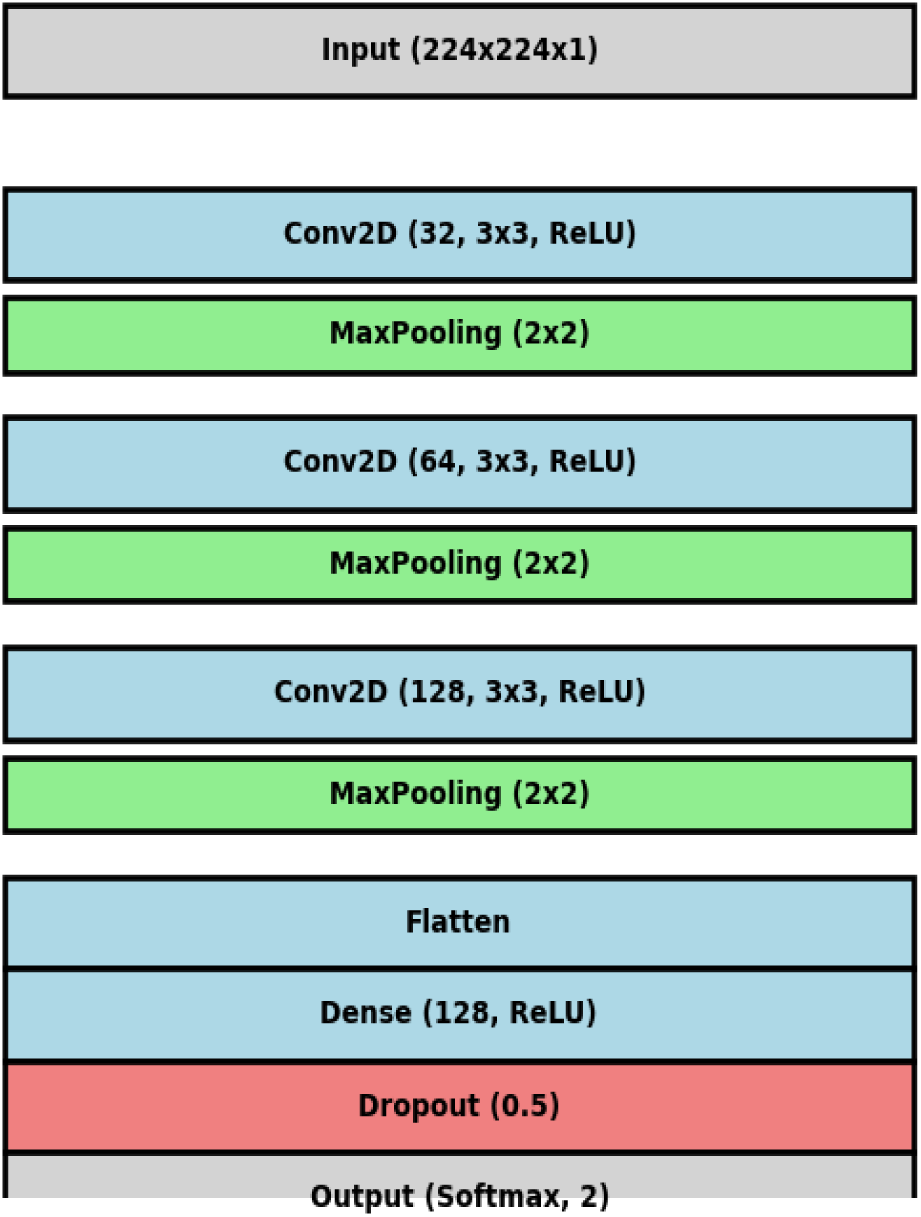
CNN model Architecture

The CNN Model summary is shown in Table 3

**Table 3.** CNN Model Summary.

| Layer (type) | Output Shape | Param # |
| --- | --- | --- |
| **conv2d (Conv2D)** | (None, 222, 222, 32) | 320 |
| **max_pooling2d (MaxPooling2D)** | (None, 111, 111, 32) | 0 |
| **conv2d_1 (Conv2D)** | (None, 109, 109, 64) | 18,496 |
| **max_pooling2d_1 (MaxPooling2D)** | (None, 54, 54, 64) | 0 |
| **conv2d_2 (Conv2D)** | (None, 52, 52, 128) | 73,856 |
| **max_pooling2d_2 (MaxPooling2D)** | (None, 26, 26, 128) | 0 |
| **flatten (Flatten)** | (None, 86528) | 0 |
| **dense (Dense)** | (None, 128) | 11,075,712 |
| **dropout (Dropout)** | (None, 128) | 0 |
| **dense_1 (Dense)** | (None, 2) | 258 |

Total params: 33,505,928 Trainable params: 11,168,642 Optimizer params: 22,337,286

**Table 4.**
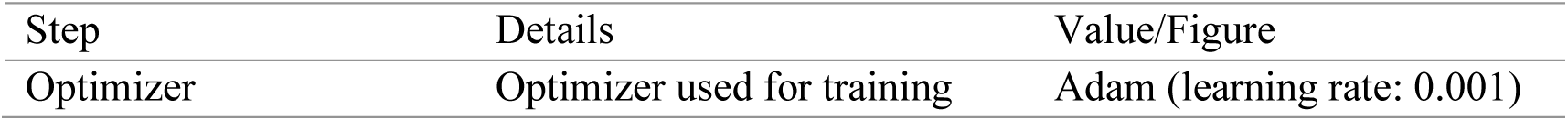
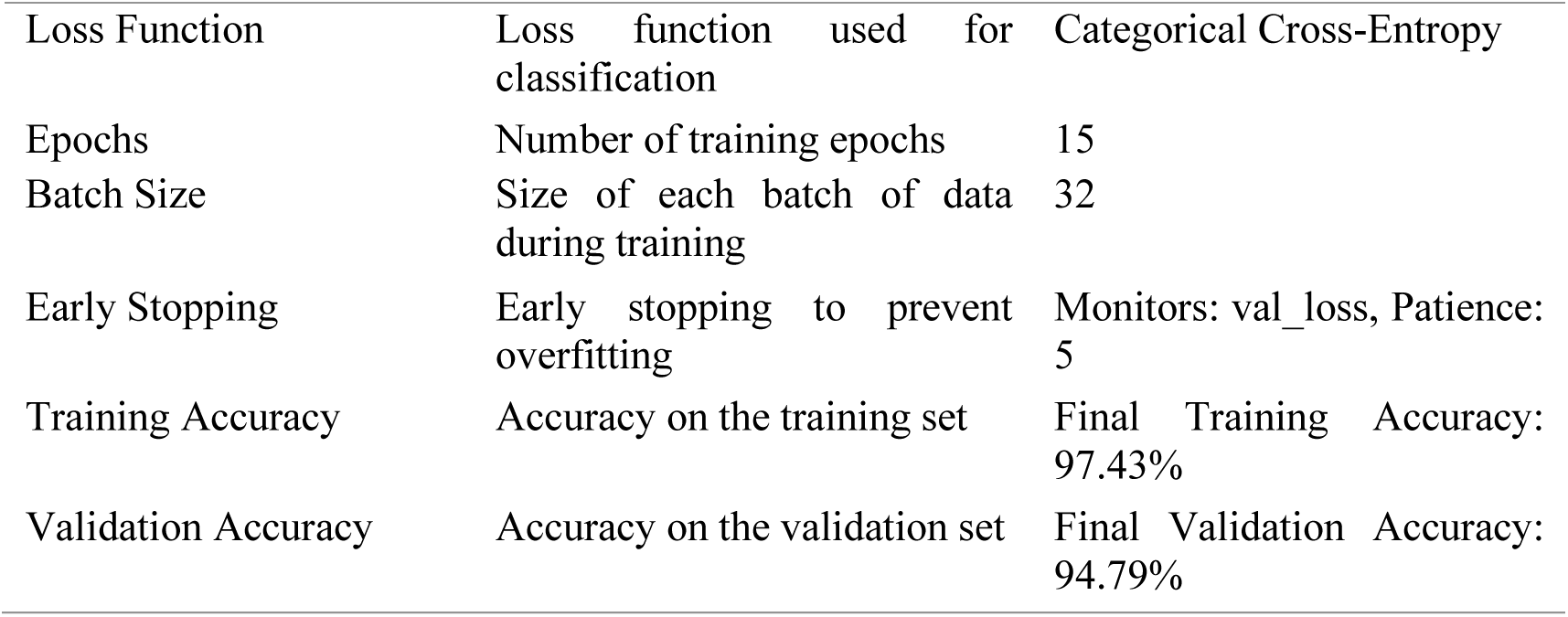
The Training Process involved in building the model.

## 4. Results

The results of this study are divided into two categories. The patient-centered system and the clinician-centered system.

### 4.1 Patient-Centered System

The results from this system include the model development, the mobile application, and the wearable sensor device developed for the testing and validation of the model.

For the model development, the model showed a very strong performance across the various train-test splits, consistently achieving high accuracy, sensitivity, specificity, and AUC values, as seen in Table 5.

**Table 5.**
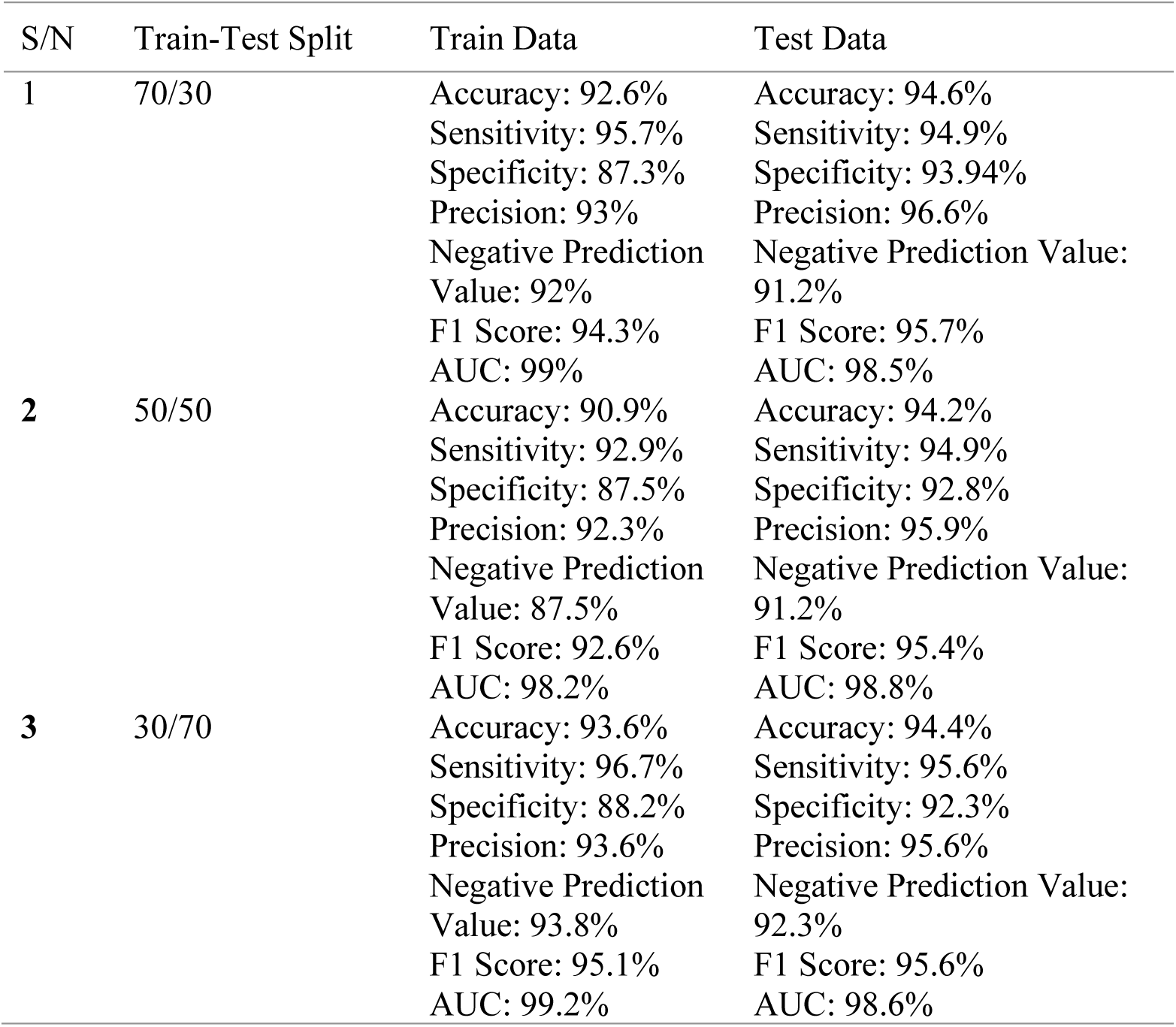
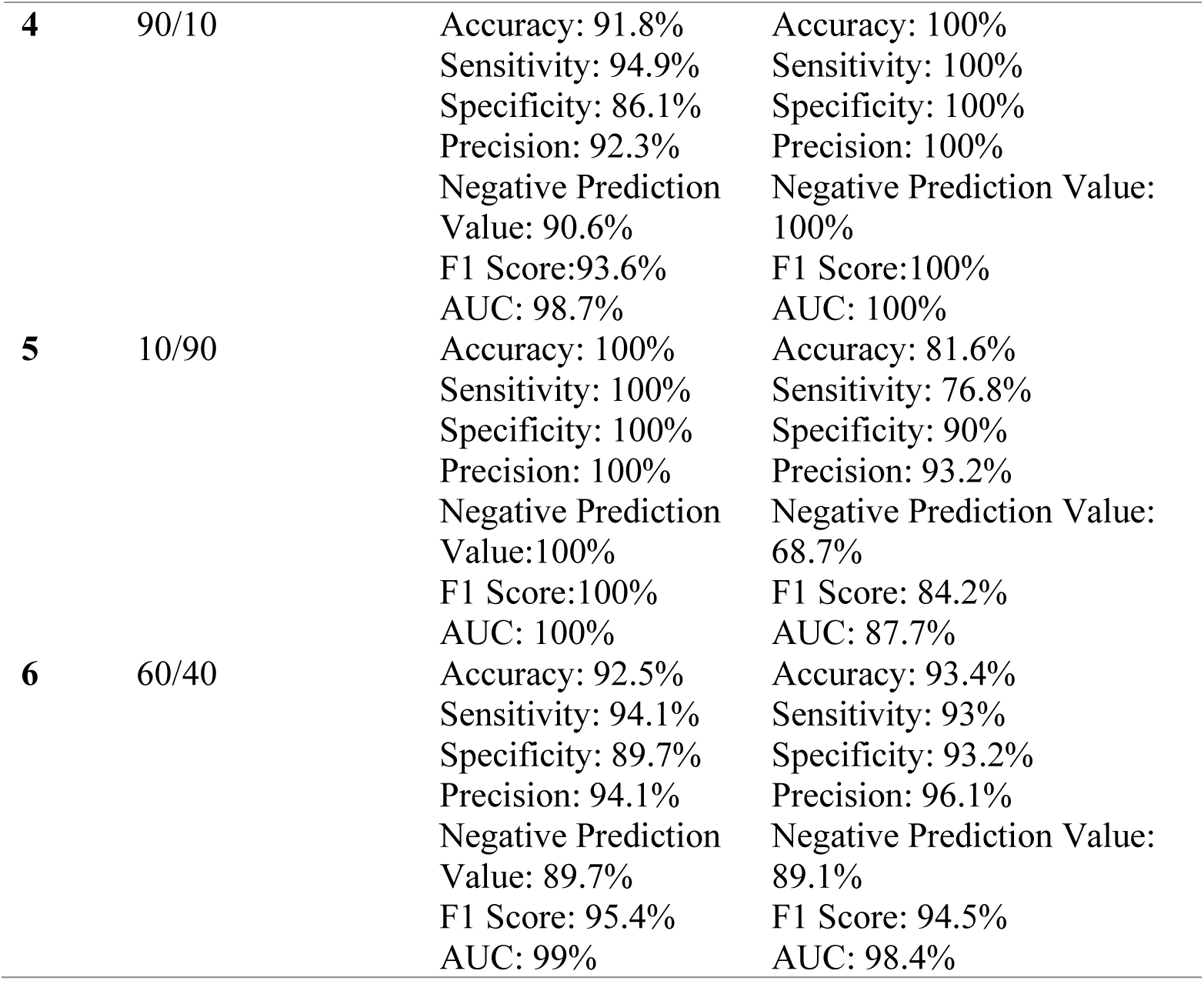
Performance Metrics of the Patient-Centered System.

The 70/30, 60/40, and 30/70 splits achieve the optimal training efficiency and generalization performance because they produce test accuracy results of 94.6%, 93.4% and 94.4% while maintaining AUC values above 98%. This indicates a good discriminatory power. The confusion matrix of the 70/30 split is shown in Fig. 4a and b

**Fig. 4a.**
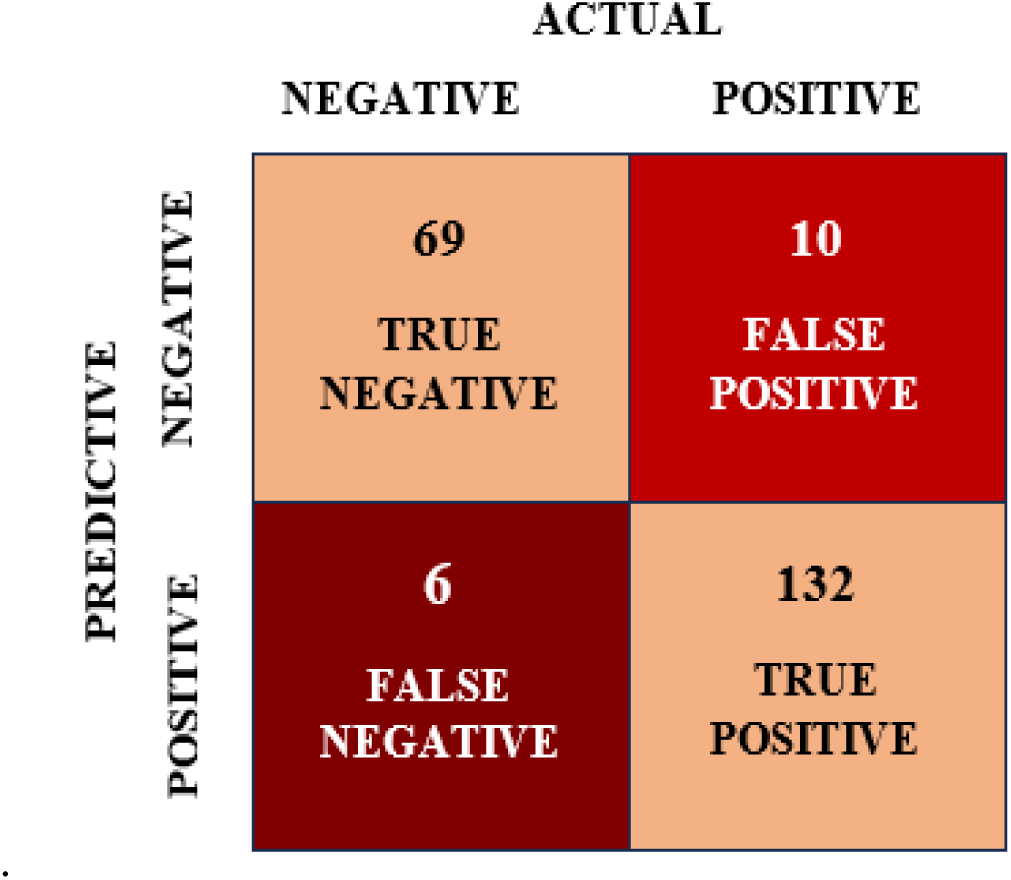
Confusion Matrix for train data

**Fig. 4b.**
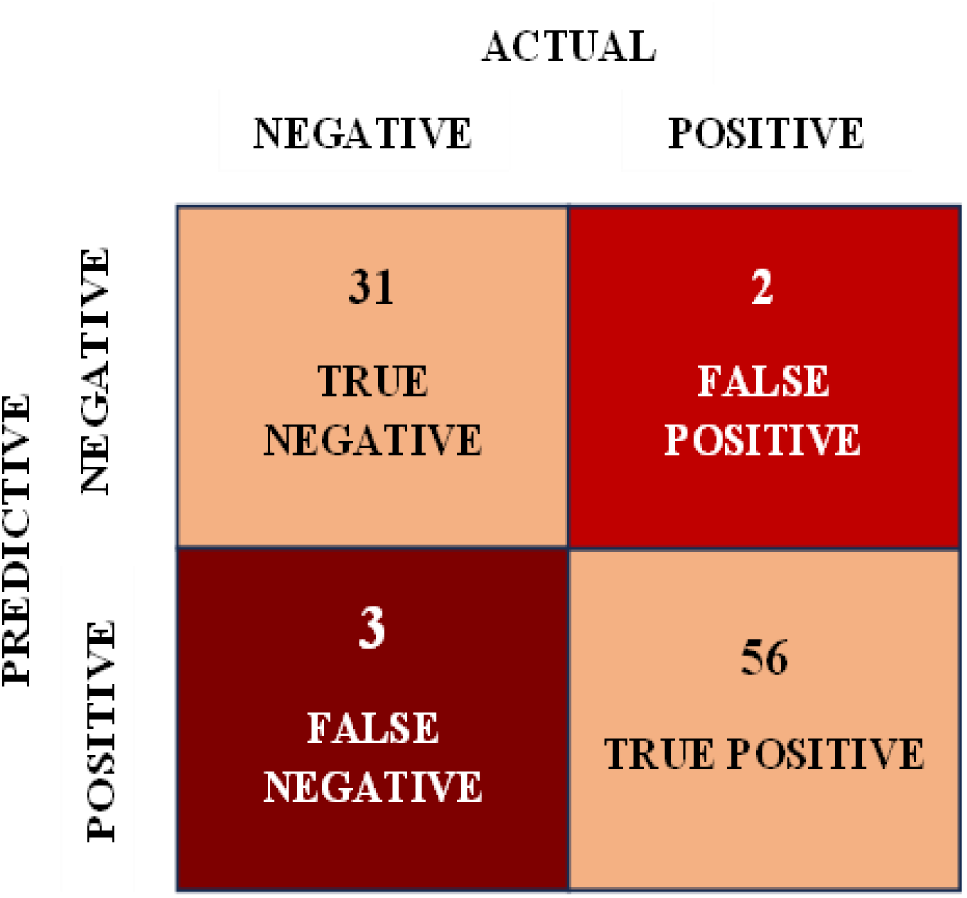
Confusion Matrix for test data

The confusion matrix of the training data provides a comprehensive assessment of the performance of the binary classification model. The matrix reveals that the model correctly predicted “No” 69 times and “Yes” 132 times, but it had 6 false negatives and 10 false positives.

The matrix reveals that the model correctly predicted “No” 31 times and “Yes” 56 times, but it had 3 false negatives and 2 false positives. The model maintains excellent stability because it correctly identifies all positive cases that belong to the “Yes” class. However, in the case of a 10% reduction (10/90) of the dataset, the model achieved a perfect training accuracy of 100% while achieving a low test accuracy of 81.6% and sensitivity of 76.8%, implying overfitting as a result of a lack of sufficient training data.

The 90/10 split, which allocated 90% of the data to training, showed a more balanced outcome, as illustrated in Fig. 5a and 5b.

**Fig. 5a.**
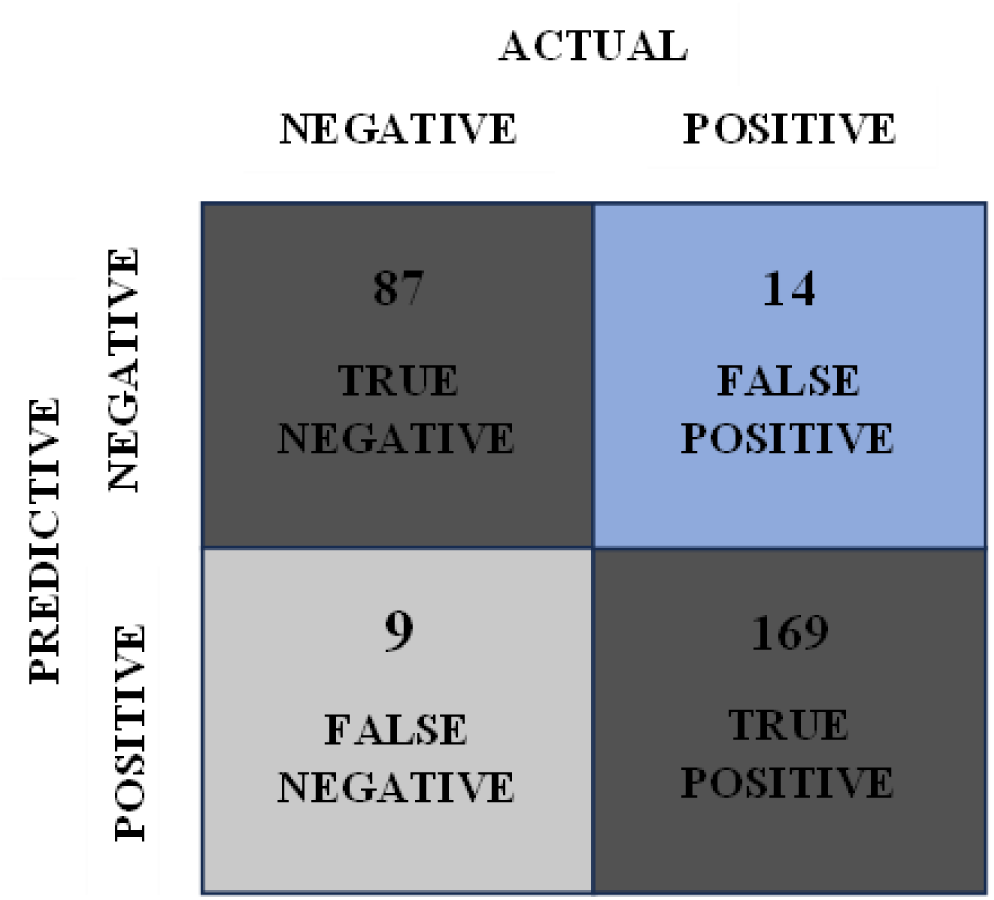
Confusion matrix for train data

**Fig. 5b.**
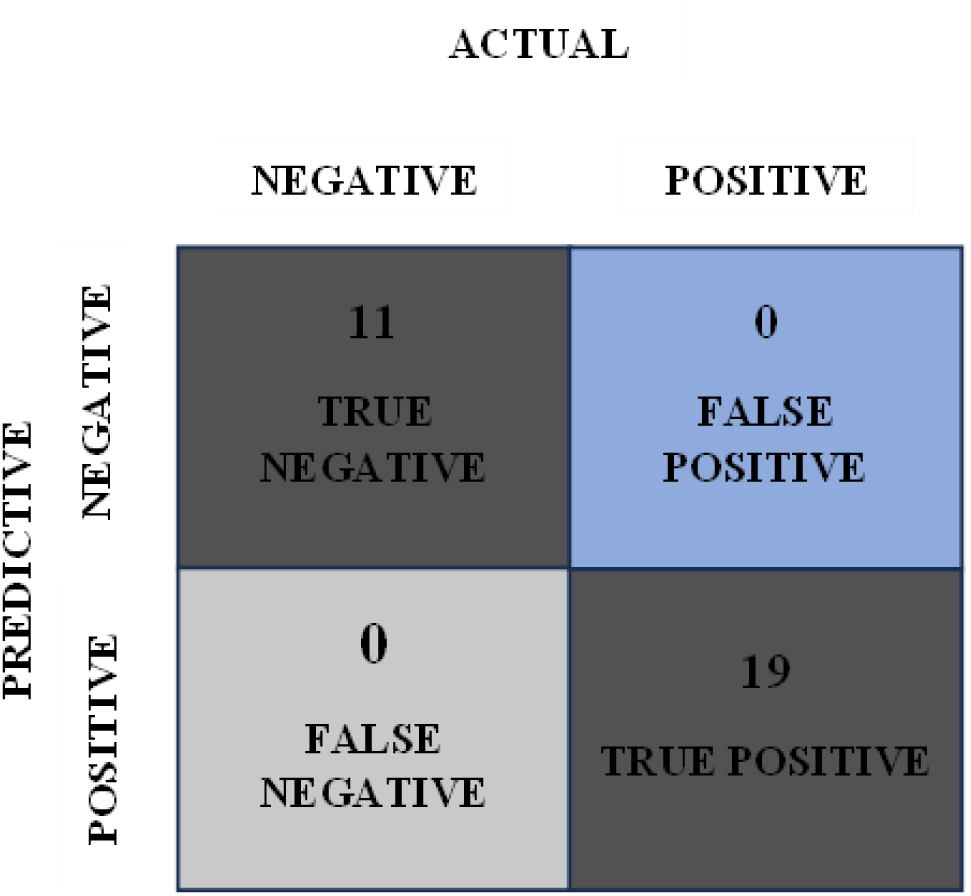
Confusion Matrix for test data

For the training data, the matrix reveals that the model correctly predicted “No” 87 times and “Yes” 169 times, but it had 9 false negatives and 14 false positives.

The metrics reveal that the model correctly predicted “No” 11 times and “Yes” 19 times, with no false negatives and no false positives, reflecting perfect classification on the small test set for this split.

The mobile application development produced a successful product, which featured an easy-to-use graphical user interface (GUI) that allowed users to access all features without difficulty. Similarly, the wearable sensor device was completed successfully. As shown in Fig. 6, the device integrates a temperature sensor alongside heart rate and oxygen saturation monitoring, housed in a wrist strap design for user comfort. Testing confirmed that the device operated correctly, successfully monitoring and recording data across all three vital signs.

**Fig. 6.**
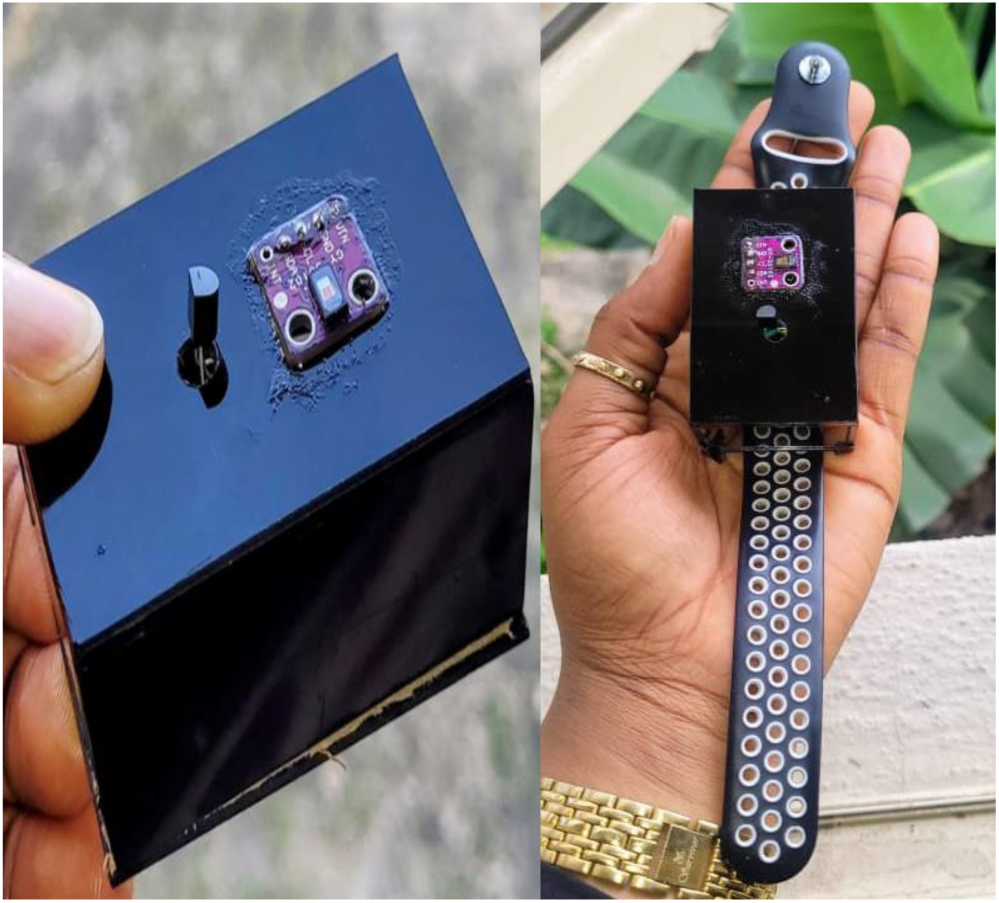
Wearable sensor device

**Fig. 7.**
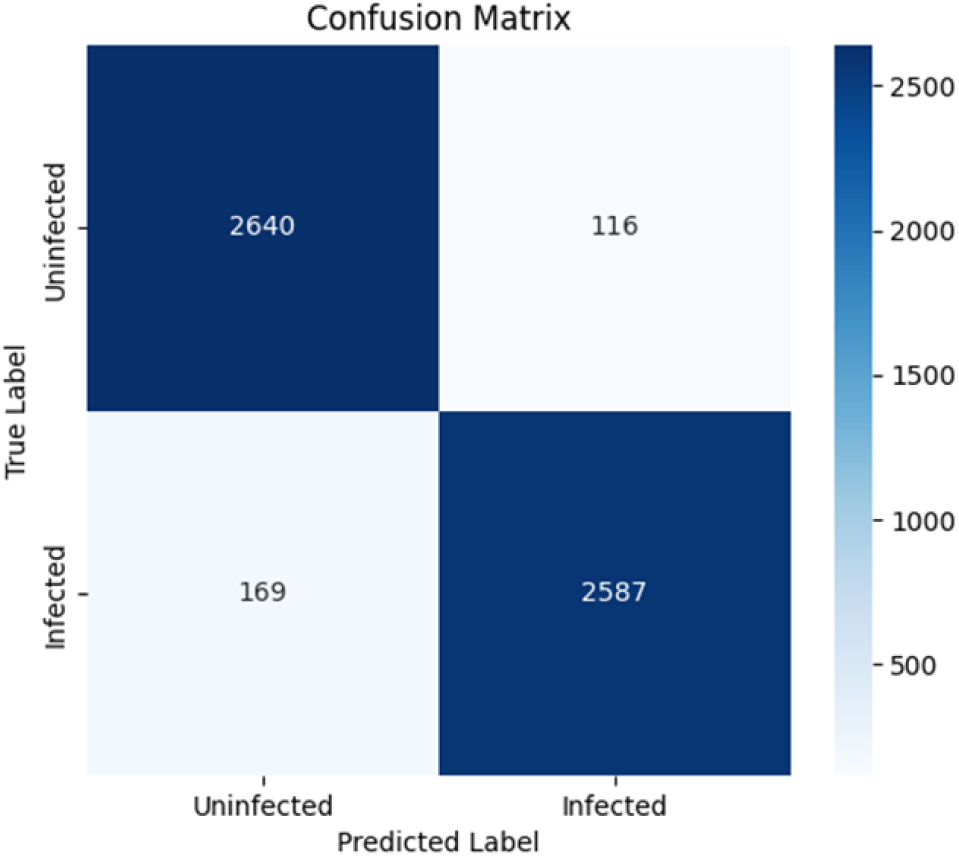
Confusion Matrix of the CNN Model

### 4.2 Clinician-Centered System

Evaluation metrics were used to assess the performance of the malaria CNN model. The evaluation assessed four essential performance metrics, which consist of accuracy, precision, recall, and F1-score.

The confusion Matrix of the model reveals that the CNN model accurately recognized 2,587 true positive cases (TP) and 2,640 true negative cases (TN), while 116 false positives (FP) and 169 false negatives (FN) were noted. From these confusion matrices, we can calculate standard performance metrics like recall, sensitivity, and accuracy. The formulas for calculating them are as follows.

Accuracy: This measures the overall accuracy of the model.

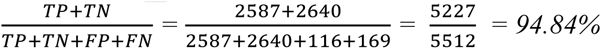

Sensitivity (Recall): This measures the model’s ability to correctly identify positive cases

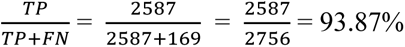

Specificity: This measures the model’s ability to correctly identify Negative cases

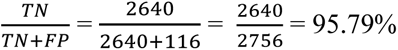

Precision (Positive Predictive Value): This measures the accuracy of positive predictions made by the model

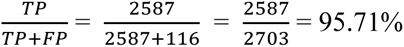

Negative Predictive Value: This measures the accuracy of Negative predictions made by the model

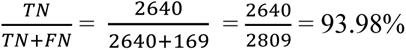

F1 Score: This measures the balance between precision and sensitivity.

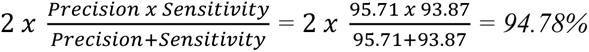

AUC: This measures the model’s ability to distinguish between parasitized and uninfected cases across all classification thresholds. Based on the ROC curve, the model achieved an AUC of 0.84, indicating strong discriminatory power.

These performance metrics speak highly of the integrity of the model’s classification ability in malaria disease prediction. A sample prediction using the model is shown in Fig. 8a & b

**Fig. 8a.**
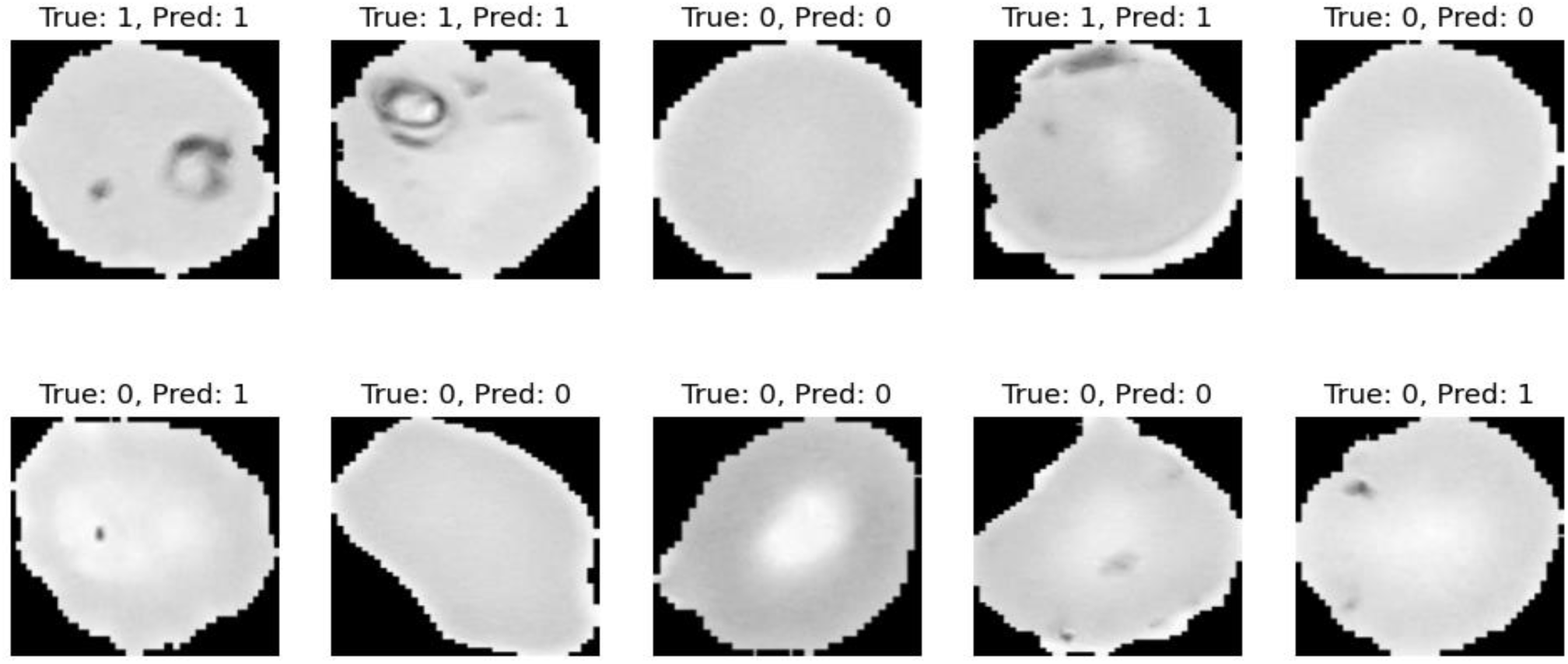
Malaria Sample Predictions

**Fig. 8b.**
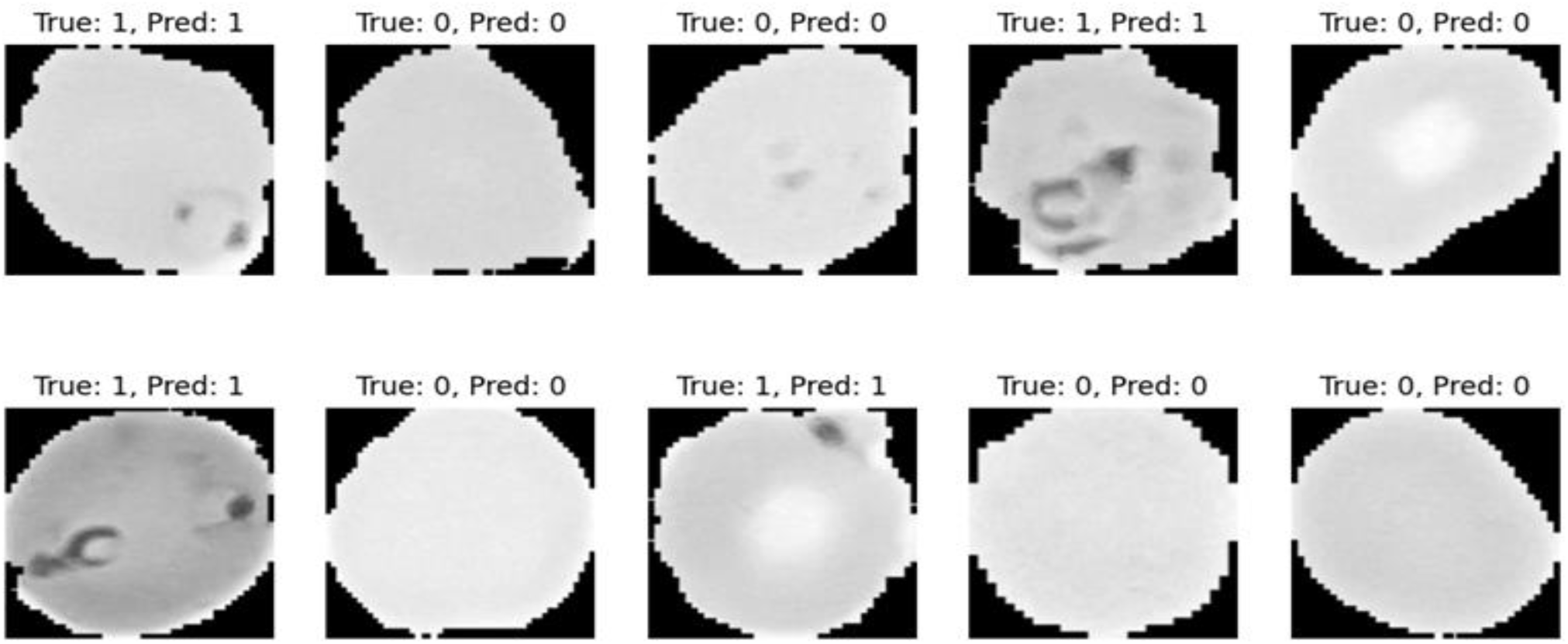
Malaria Sample Predictions

Twenty sample prediction images were examined to evaluate the model’s efficacy in more detail. The images demonstrate both right and wrong case identification through visual displays of the model’s classification output. The research findings reveal both the strengths and weaknesses of the model through its ability to detect malaria - specific features and its errors in missing or incorrectly identifying infections.

## 5. Discussion

### 5.1 Patient-Centered System

This system consists of a mobile application and a wearable sensor device. The results from the system analysis demonstrate the significance of biosignals like temperature, heart rate, and Oxygen saturation in the prediction of malaria, alongside the symptoms experienced by patients. These were very instrumental to the successful prediction of malaria and non-malaria cases. Patients with malaria showed elevated body temperature, heart rate, respiratory rate, and systolic blood pressure, along with slightly lower SPO_2_ levels, compared to patients without malaria. Of these, only the differences in temperature and heart rate were statistically significant in the multivariate model (Table 1); respiratory rate, blood pressure, and oxygen saturation trended in the expected direction but did not reach significance. The study results confirm earlier research [27, 28], which demonstrated that these essential body signs serve as essential monitoring tools for patients while validating their use as diagnostic markers.

The machine learning algorithm, Multivariate Logistic Regression, used for the learning process performed excellently, providing high accuracy levels and scores. When the model was overtrained, a higher level of accuracy, as well as other confusion matrices and statistics, were observed. When the model was poorly trained or had less data to learn from, as was seen in the 10/90 train-test split, the model did not perform as well as others. The model shows consistently strong predictive performance across different train-test splits because it achieves high accuracy and precision and maintains high sensitivity and specificity. The model also shows reliable performance because it achieves similar results across different splits, which indicates its ability to predict new data with high accuracy. Similar results have been seen in previous studies. A penalized logistic regression model to predict Plasmodium falciparum (Pf) antigen positivity based on clinical, environmental, and sociodemographic factors achieved an AUC of 84% on the training set and 83% on the test set [29]. These findings emphasize the importance of clinical and environmental factors in malaria prediction, which is consistent with our study’s approach of integrating biosignal and symptom-based assessments.

The success of this model provides a seamless integration into the developed mobile application, Malacare, and the wearable sensor device. The developed wearable sensor successfully recorded temperature, Heart rate, and Oxygen Saturation levels when tested. It also transmitted the results to Malacare, which successfully carried out patient symptoms, as well as sociodemographic assessment, and received signal details from the wearable sensor device. With these details, it successfully predicted the outcome of malaria. This technique is in line with a wearable, Internet of Things-based e-health monitoring device for assessing vital signs in real time, such as heart rate, oxygen saturation, and body temperature. Their method effectively moved patient data to a cloud-based platform, allowing for expert diagnosis and remote health monitoring. The study demonstrated remarkable accuracy levels, with heart rate and SpO₂ values above 96% accuracy when compared to typical smartwatch measurements [30]. These findings validate the use of wearable technology in remote health evaluations and support the integration of biosignal-based malaria prediction into our system.

The results of this Malaria Pre-Screening Technology using Artificial Intelligence present a new solution to the burgeoning problems of malaria, bringing the diagnosis closer to the patient. This way, a pre-assessment could be done very fast and in a convenient way, and also help patients gain insights early enough before seeking medical attention. Furthermore, the system provides essential benefits to medical facilities that operate with minimal patient-to-doctor staffing ratios. The patient-centered system begins with symptom and biosignal analysis to generate an initial diagnosis, but requires clinical evaluation to establish the final diagnosis. Medical image analysis serves as a vital element of the clinician-centered system, which works to enhance diagnostic precision. However, the model’s development on a relatively small sample (n=309) and its evaluation without an external validation cohort limit the generalizability of these findings to broader patient populations.

### 5.2. Clinician-Centered System

This system is made up of a custom CNN model that is deployable for the analysis of cell images and the prediction of parasitized and uninfected malaria cases. From the performance analysis, the model was able to perform very well in separating malaria-positive from uninfected cases with a precision of 95.71%, implying a low level of false positives and hence the minimal probability of misclassifying healthy cases as malaria-positive. In addition, the model had a recall of 93.87%, and therefore it was able to identify most of the infected cases and avoid the chances of missing the malaria patients.

Furthermore, the model’s F1-score of 94.78% demonstrates its operational reliability and suitability for real-world diagnostic systems. These results are consistent with findings where a deep neural network achieved a classification accuracy of 93.1% in identifying Plasmodium falciparum-infected cells, further supporting the model’s strong generalization ability when applied to malaria microscopy images [31].

However, the model has certain drawbacks. There is potential for improvement, as a small number of infected cases were still incorrectly labelled as uninfected. The model could achieve improved pattern recognition through hyperparameter optimization and data augmentation methods [32], which would result in lower error rates. The evaluation of its performance on external datasets can also show its effectiveness with different population groups.

Looking ahead, future research could explore explainable AI, ensemble learning, or hybrid architectures to boost detection accuracy and strengthen the model’s overall reliability [33]

## 6. Conclusion

This research study contributes significantly to the prescreening of malaria among patients. In particular, the system provides patients and clinicians with an efficient and convenient way of diagnosing malaria through a multimodal and synergetic combination of artificial intelligence and the clinical examination of the symptoms, signs, and image-based prediction. The AI models performed efficiently during the training and testing processes, proving their ability to effectively differentiate between malaria-positive and uninfected patients. This work represents a meaningful step forward in malaria detection, integrating AI-based symptom evaluation with deep learning-based image analysis to support real-time patient assessment within an intelligent detection system. The approach shows promise for identifying malaria at its earliest stages, enabling clinicians to select appropriate treatment options and contributing to broader efforts toward global malaria eradication.

## Data Availability

All data produced in the present work are available online and within the supplementary materials contained in the manuscript.

https://www.kaggle.com/datasets/iarunava/cell-images-for-detecting-malaria

## Declarations

No funding was received for conducting this study.

### Ethics Declaration

This study used publicly available, de-identified datasets. As the data were fully anonymized and publicly accessible before this study, no additional ethical approval was required.

